# An end-to-end workflow for statistical analysis and inference of large-scale biomedical datasets

**DOI:** 10.1101/2020.01.09.20017095

**Authors:** Elyas Heidari, Mohammad Amin Sadeghi, Vahid Balazadeh-Meresht, Nastaran Ahmadi, Mahmoud Sadr, Ali Sharifi-Zarchi, Masoud Mirzaei

## Abstract

Throughout time, as medical and epidemiological studies have grown larger in scale, the challenges associated with extracting useful and relevant information from these data has mounted. General health surveys provide a good example for such studies as they usually cover large populations and are conducted throughout long periods in multiple locations. The challenges associated with interpreting the results of such studies include: the presence of both categorical and continuous variables and the need to compare them within a single statistical framework; the presence of variations in data resulting from the technical limitations in data collection; the danger of selection and information biases in hypothesis-directed study design and implementation; and the complete inadequacy of p values in identifying significant relationships. As a solution to these challenges, we propose an end-to-end analysis workflow using the MUltivariate analysis and VISualization (MUVIS) package within R statistical software. MUVIS consists of a comprehensive set of statistical tools that follow the basic tenet of unbiased exploration of associations within a dataset. We validate its performance by applying MUVIS to data from the Yazd Health Study (YaHS). YaHS is a prospective cohort study consisting of a general health survey of more than 30 health-related measurements and a questionnaire with over 300 questions acquired from 10050 participants. Given the nature of the YaHS dataset, most of the identified associations are corroborated by a large body of medical literature. Nevertheless, some more interesting and less investigated connections were also found which are presented here. We conclude that MUVIS provides a robust statistical framework for extraction of useful and relevant information from medical datasets and their visualization in easily comprehensible ways.

## 1 Introduction

In a time where copious amounts of data are being gathered in all areas of medicine, the ability to extract valid and relevant information from these large datasets is paramount. The usual practice for analyzing multivariate data includes three general steps: I. pre-processing and quality control, including identification and exclusion of outliers and low-quality samples and imputation of missing data; II. multivariate analysis using a variety of approaches, including hypothesis testing, predictive models, correlation analysis, graphical modeling, etc.; and III. visualization and interpretation of the results, including uni-, bi- and multivariate plots, interactive and dynamic graphical representations, network visualization of interactions, etc. Several R packages have been developed so far for carrying out any of the above tasks. However, since there is no single package providing all of these functionalities, conducting the complete analysis requires using many different packages and re-adaptation of data among them, which is a cumbersome challenge for many people of different scientific backgrounds who need to analyze their multivariate data. Datasets acquired from health surveys are a good example of such data, the challenges they present, and the shortcomings of routine statistics in tackling these challenges:

1. These studies usually survey a statistically diverse range (i.e., both categorical and continuous) of variables. Traditional statistics would require using different statistical tests based on the type of comparison (e.g., categorical vs. categorical or continuous vs. categorical) being studied. However, if it is intended to extract the most important relationships from the dataset as a whole, it will prove difficult to compare findings across the boundaries of such different statistical tests.
2. These studies frequently require multiple individuals in multiple centers to collect the data. Therefore, the datasets resulting from these studies must undergo a rigorous quality control process. As will be explained in the sections on data preprocessing and population structure, methods that are superior to the current conventional tools are available for performing this quality control process.
3. While analysis of medical datasets, small and large, is a routine undertaking, these studies are usually based on and guided by some previous hypotheses. In other instances, only a small number of hypotheses are tested on a limited number of variables in large datasets. Such reductionist approaches make the research process prone to both selection (i.e., how the source for data is determined) and information (i.e., how the data is gathered and interpreted) biases. Furthermore, any inference in such studies may be subject to various confounding factors and it becomes impossible to define a global structure of interactions between the variables.
4. The large size of such datasets renders the traditional analytic methods of regression analysis, correlation analysis, and hypothesis testing quite inadequate for extracting meaningful results. As an example, it is well known that tests for statistical significance lose relevance in large datasets, giving very significant p-values for the faintest of differences [1]. As a result, other measures need to be taken into consideration when comparing the identified relationships, one example being effect sizes for defining the strengths of the relationships.
5. As explained more comprehensively in the methods section, traditional methods for predictive model construction such as those based on linear models lose their accuracy in large datasets of high dimensions.
6. The shear amount of such datasets require any method designed for global analysis to not only be statistically robust, but also computationally efficient.

As a solution to these challenges, we propose an end-to-end analysis workflow using the MUVIS (MUltivariate analysis and VISualization) package in R statistical software [2]. Our focus in designing this workflow is to use robust statistical analyses for extracting useful biomedical information from large datasets and presenting the results in visually simple and readily understandable ways. MUVIS is a toolkit for analyzing multivariate datasets through the following general steps (Figure 1). The pertinent details of each method and how they should be interpreted are expanded on later in the manuscript. Furthermore, a mathematically intensive description of the methods is given in Supplementary file 1.

**Figure 1.**
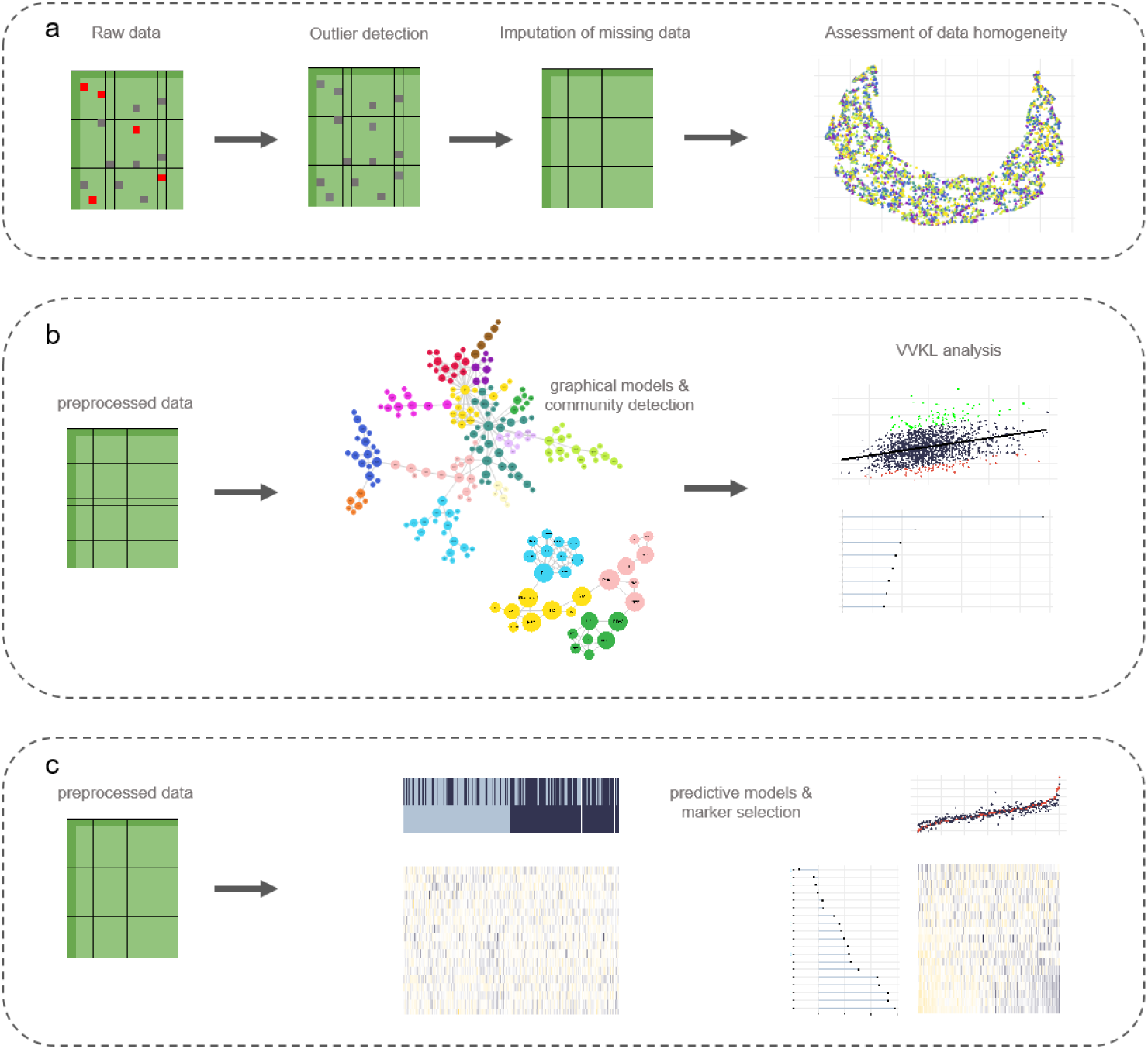
The general workflow of dataset analysis using the MUVIS package. **a)** The first phase of the workflow preprocesses the data in order to prepare it for further analysis. This phase consists of outlier detection, replacement of outliers and missing data through imputation, and assessment of data heterogeneity. **b)** The second step consists of unbiased exploration of the dataset for identifying relationships between variables through graphical models. These relationships are further studied using the VVKL method. **c)** Finally, the dataset is used for constructing prediction variables using the Elastic Net algorithm.

We apply these analytical methods on a dataset acquired from the recruitment phase of Yazd Health Study (YaHS), a population-based cohort study of 10050 individuals aged uniformly from 20 to 70 years, conducted in November 2014 in the Greater Yazd Area of Iran [3]. The YaHS study uses a verified questionnaire of close to 300 questions that are collected by trained interviewers. The questions regard: a) demographics, b) physical activity, c) sleep quality and quantity, d) mental health, e) past medical history (PMH) of chronic disease and surgical operations, f) dental health, g) history of accidents, h) dietary habits, i) occupation and social life, j) use of traditional medicine, k) smoking habits and drug addiction, l) women’s health, and m) quality of life. Blood pressure, pulse rate, anthropometrics, and body composition indices were recorded for all participants. Moreover, biochemical and hematological indices were measured in blood samples from a subset of 4010 participants.

In addition to explaining the application of the MUVIS workflow, we report some of the more noteworthy findings on this subset of participants from the YaHS dataset.

## 2 Methods

### 2.1 Data preprocessing

In this phase, an inbuilt function in the MUVIS package for outlier detection and replacement with imputed data was applied to the dataset. The outliers are detected using an anomaly detection algorithm described by Vallis et al. [4]. This method orders data points of the variable of interest in an increasing manner and identifies both the global trend and piecewise local trends of these data points. Then, the algorithm uses statistically robust methods (based on piecewise medians) to find the outliers. This algorithm is more sensitive than the usual Interquartile range-based outlier detection methods in that it can also detect instances where the data does not follow a smooth trend and shows a multimodal distribution. Following the detections of these outliers, they are excluded from the data and replaced by imputed data through mean (for continuous variables) and mode (for categorical variables) functions. These data points comprised nearly 2.8% of the YaHS dataset. After this preprocessing phase, the data has 34 continuous and 181 categorical variables.

### 2.2 Population structure

The data from large health surveys is usually collected in multiple sites, by multiple individuals, and during multiple time frames. In such instances, devising clear standards for data collection and harmonization are paramount. Nevertheless, it may be observed that within- and between-site/time frame distributions of data differ, indicating the presence of confounding factors such as the time and location where the data was gathered. In any data analysis pipeline, it is important to rule out such variations as they may obscure the actual relationships between data variables.

Dimension reduction techniques are useful tools in visualizing complex high dimensional data in simplified ways. Such techniques transform the data in such a way that they could be visualized using two dimensions while encapsulating the global distribution of the original data. This visualization enables examination of data heterogeneity (i.e., aggregation of data points into multiple clusters), homogeneity (i.e., uniform distribution of data points across the dimensions), and, if heterogeneous, identification of the underlying factors aggregating the data points (e.g., time and site of data collection). To this end, we propose using a dimension reduction technique called Uniform Manifold Approximation and Projection (UMAP). UMAP is superior to the more commonly used principal component analysis (PCA) in that it is better in capturing the relationships in non-linear data structures; i.e., high-dimensional data that are not sequential. In the YaHS dataset, we try to capture the population structure by reducing the data dimensions from 34 (number of laboratory measurements) to 2. As indicated in Figure 2a, the data points are uniformly distributed throughout the dimensions and, hence, are homogeneous. Furthermore, gender and age, two important confounders on any observed relationship, are uniformly distributed in our study population Figure 2b).

**Figure 2.**
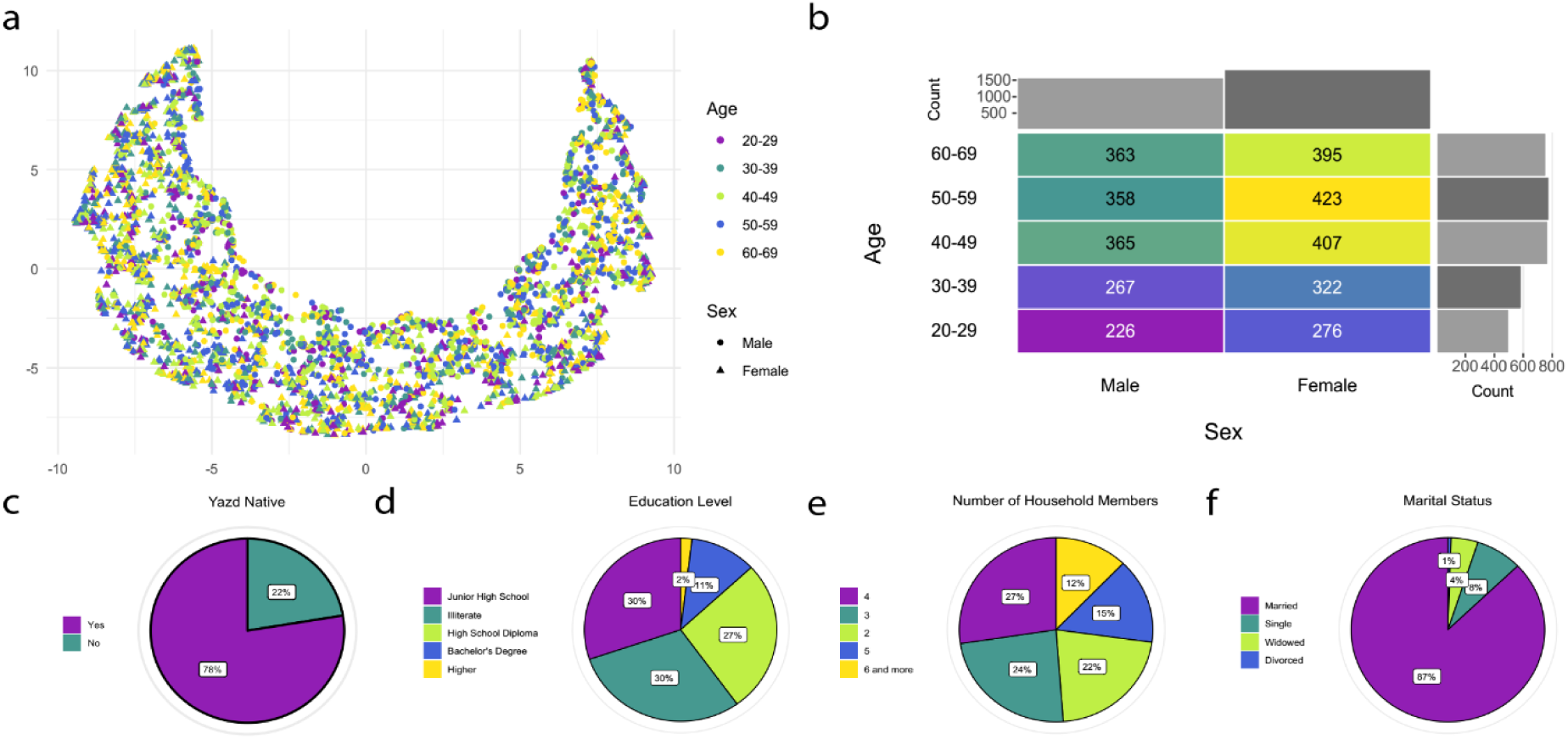
Population overview. **a)** UMAP plot of the population. The population is projected on 2 dimensions with UMAP; each point represents a sample from the population. The color and shape of each point corresponds to age group and gender. **b)** Distribution of gender and age in the population. The number in each cell represents the number of samples within that specific combination of gender and age group. **c-f)** Pie charts of location of birth **(c)**, education level **(d)**, number of household members **(e)**, and marital status **(f)**.

### 2.3 Probabilistic Graphical Models (PGMs)

Once the data is preprocessed, the first step in the analysis pipeline is unbiased exploration of all possible relationships among variables using graphical models. These models identify interactions (relations) between variables and visualize them as interaction networks. It is important to note that these graphs do not visualize all of the identified relationships between variables since that would construct networks with too many relationships to interpret. Instead, these models focus on only the most significant relationships and, therefore, build sparse networks. Two main algorithms for graphical models, each having multiple methods of model selection for constructing sparse networks (i.e., determining the relationships that should be included in the graph) are included in the MUVIS package.

In addition to the relationships networks, PGMs also yields another line of insight. In a PGM, each node may be viewed as containing or, in other words, sharing information about its adjacent nodes, representing a process of “information flow”, also referred to as “influence flow”, between them [5]. For each pair of nodes, a shortest path for this information flow based on the number of edges could be calculated. For each node, betweenness centrality is then defined as the number of these shortest paths that pass through the node. Therefore, it could be stated that nodes with the highest betweenness centrality in the model have the most significant control on the flow of information within the whole graph.

#### 2.3.1 Gaussian graphical model of continuous variables

A Gaussian graphical model (GGM) is used to construct the graph of associations among continuous variables in which each node represents a Gaussian random variable, and each edge demonstrates a non-zero estimated partial correlation between two variables. The size of each node is relative to the number of edges it connects to. In essence, GGM assumes normality of data and treats variables as Gaussian random variables. This assumption leads the variables to have a sparse (zero-dense) inverse of a covariance matrix, such as S, in which each value S_ij_ can be shown to represent the partial covariance (or equivalently, partial correlation) between variables X_i_ and X_j_. It can also be shown that zero partial correlation between two Gaussian variables implies their conditional independency [6]. Conditional independency (dependency) refers to independency (dependency) of two variables given all other variables, and likewise partial correlation is defined as the correlation coefficient of two variables given all other variables. Thus, modeling the inverse of the covariance matrix with a graph, in which each node represents a variable and each edge represents a non-zero partial correlation, given the assumption of normality, leads to a sparse graph like the one shown in Figure 3. Therefore, in such a graph, each edge indicates a significant relation between two variables in the sense that there is no other variable describing the relation between these two variables. Importantly, the word significance here does not necessarily convey the classical concept of statistical significance and its exact meaning depends on the model selection method which has been used for constructing the GGM.

Here we use the graphical lasso (glasso) algorithm in order to construct our GGM. The objective of glasso is to estimate the inverse of the covariance matrix in a penalized setting using L1-norm which leads to a sparse estimation [7]. Following the construction of the GGM, we use a community detection algorithm using the Louvain method to find the communities of highly related variables within the graph [8]. Briefly, this algorithm finds groups of variables that are highly interconnected and form edge-dense clusters. The validity of the model could then be assessed based on how variables are clustered into different communities.

#### 2.3.2 Minimal Forest

Unlike the GGM, the minimal forest algorithm could be used for estimating relations among both continuous and categorical variables. In addition, the MF is much more robust and faster in dealing with datasets with large (tens to hundreds) numbers of variables [9]. Since this algorithm analyses categorical variables as well continuous ones, partial correlation may not be used for identification of relationships. Instead, the associations in the minimal forest are determined based on a concept called mutual information.

Mathematically, mutual information is the Kullback-Leibler divergence (explained later) between the joint probability distributions of two variables and the product of the independent ones. In simple terms, the more the information shared between two variables, the higher their mutual information. As was the case in the GGM, each node represents a variable, and each edge represents the association between two nodes. While the exact resulting graph would differ based on the model selection method used (Supplementary file 1), the algorithm generally proceeds to connect all nodes with the minimum number of edges (minimum complexity); therefore, only the edges with the biggest weights remain in the final graph and represent the most important connections of the variables. Here we use the version of the algorithm proposed by Abreu et al. [10], where the objective is to maximize the sum of mutual information values on existing edges subtracted by a linear function of network complexity (e.g., number of edges), thus, leading to a sparse graph.

After the graph is constructed, a community detection algorithm is used to find clusters of highly related variables similarly to the GGM.

### 2.4 Violating Variable-wise Kullback-Leibler Divergence (VVKL)

The relationships identified by the previous methods all require further exploration using conventional statistical methods to determine the nature of the relationship. For associations between continuous variables, such as those identified by the GGM, the method of choice has commonly been the construction of scatter plots and calculation of correlation coefficients. We believe, however, such scatter plots are able to yield much greater information. Since the correlation between two variables is usually far from perfect, some data points could be found which do not conform to the general trend; i.e., show higher or lower values than expected compared to the trend predicted by the correlation coefficient.

In this workflow, we suggest applying the Violating Variable-wise Kullback-Leibler divergence (VVKL) method to study the possible causes behind these observed deviations. Mathematically, KL divergence is a measure of the difference between two distributions.

The VKL uses KL divergence to find and rank all variables which differ the most between different states of the studied condition. In the VVKL method, we compare two groups of data points: a) Those with the highest upward deviation from the general trend; and b) those with the highest downward deviation from the general trend. The VKL method is applied to identify variables that are differently distributed between the two groups. An example VVKL analysis of the relationship between blood cholesterol and HDL levels is presented below. The power of the VVKL lies in the fact that it readily ranks the identified influencing factors, whether categorical or continuous, based on the calculated KL divergence. This KL divergence acts essentially as the effect size of the influence exerted by the identified variables and is comparable between both types of variables.

### 2.5 Elastic Net

Once the relationships have been identified, they could be utilized for constructing models to predict a medical condition of interest. While multivariate linear regression has been the method of choice for building such models, when the number of predictors becomes too large as in our dataset, the variances of the estimates and hence the total error of the model increases substantially [11]. Therefore, linear models tend to fail in datasets with a large number of variables. A better method for such prediction, however, can be applied to the whole dataset and utilize all its variables as predictors. To this end, one may use regularized linear models such as LASSO, which imposes an L1-norm penalty; Ridge regression, which imposes an L2-norm penalty; or Elastic Net, which imposes a linear combination of L1- and L2-norm penalties. Here we use the Elastic Net which is known to be superior to the other methods mentioned above, especially in settings with assumptions of both sparsity (i.e. assuming there are just a few variables that are significantly relevant to a condition of interest) and lack of complete independency between variables. In other words, Elastic Net can be shown to select correlated groups of relevant variables [12]. In order to find the best penalizing parameters of the model, we performed a 10-fold cross validation on a complete range of parameters covering both LASSO and Ridge regression.

Using the Elastic Net algorithm, one is able to construct a multivariate logistic regression model in a way that the variance and total model error are kept to a minimum. Briefly, this algorithm simultaneously forces some of the coefficients to zero (thus excluding these variables from the model) and other coefficients close to zero. Thereby, this algorithm reduces the number of predictor variables while keeping total model error low. Furthermore, the algorithm ranks the variables based on their coefficients. The number of variables yielded by the algorithm are easily determined by the researcher. As a result, this algorithms is very powerful in predicting medical conditions (categorical variables - Figure 6a) and indices (continuous variables - Figure 6b) when given a large dataset.

## 3 Results

### 3.1 Identification and exploration of relationships

#### 3.1.1 Gaussian graphical model of continuous variables

As illustrated in Figure 3, the GGMs are decomposed into four communities each. Due to the obvious confounding effect of gender on many of the variables analyzed with the GGM, one GGM has been constructed for each gender. In both graphs, community 1 clusters all variables related to anthropometric, body composition, and body fat distribution measurements alongside resting metabolic rate (MetaP). Community 2 contains all variables related to biochemical lab tests of the blood alongside measurements of blood pressure. In addition, all variables related to hematological indices are clustered into communities 3 and 4. Interestingly, the models show a difference between males and females in whether platelet-related indices are clustered alongside red or white blood cell indices. Indeed, all the communities and almost all of the relationships identified by the GGM align with our most basic knowledge of medicine. Nevertheless, some interesting and less investigated connections may also be extracted from the graphs which are presented in Table 1.

**Table 1.** Interesting findings from GGM analysis of the YaHS dataset. The findings are presented in no particular order.

| Interesting connections and findings | Previous mentions in medical literature |
| --- | --- |
| Serum uric acid concentration (UAC) and weight in males | [13] |
| UAC and fasting serum glucose (FSG) in males | [14] |
| UAC and serum triglyceride level (TG) in males and females | [14] |
| Serum total cholesterol level (Chol) and platelet number (Platelet) in males | [15] |
| UAC with Fat and BMI in females | [13, 14] |
| UAC and NeckCircum | [16] |
| Differential clustering of hematological indices between males and females | No previous mentions to our knowledge. |

**Figure 1.**
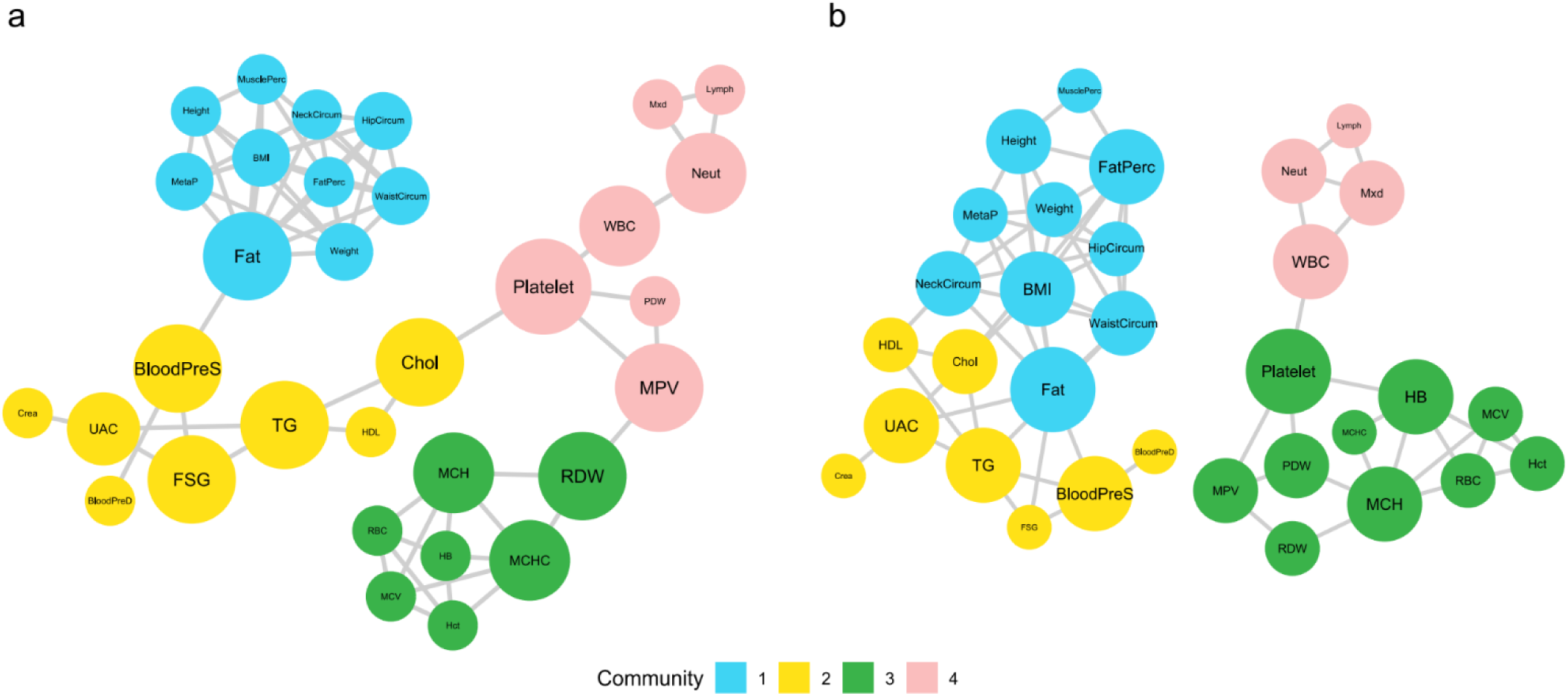
GGM of the relationships between continuous variables in males **(a)** and females **(b)**. Each node represents a continuous variable and each edge indicates a non-zero partial correlation between a pair of variables. Nodes are colored based on their community (edge-dense clusters of interrelated variables) and their size indicates the logarithm of their betweenness centrality.

In addition to the structure of the communities and individual connections between pairs of variables, it may be useful to study the nodes in the GGM based on their betweenness centrality (BC) values. In Figure 2, nodes in the GGM for males are larger due to all of them being connected to each other (Figure 2a), which is not the case for females (Figure 2b). This results in higher degrees of BC for the nodes and, hence, the larger diameters in the graph. The nodes with the highest BC and their BC values are presented in Table S1 (Supplementary file 2). In both graphs, blood platelet count (Platelet), blood triglycerides (TG), and total body fat mass (Fat) rank highly in terms of BC. However, other variables are ranked more differently between the two genders. For example, white blood cell count (WBC), blood hemoglobin (HB), blood uric acid concentration (UAC), and body fat percentage are ranked higher in females while systolic blood pressure (BloodPreS), fasting blood sugar (FSG), mean platelet volume (MPV), and red blood cell width distribution (RDW) are ranked higher in males.

#### 3.1.2 Minimal forest of all variables

Application of the minimal forest algorithm on the YaHS dataset results in a graph with 14 communities (Figure 4). The aggregation of variables into communities may hold interesting information. For example, there seems to be a general clustering of individuals into healthy/health-conscious (Communities 7 and 8) and unhealthy/health-indifferent (community 9) groups. Community 7 aggregates [higher] usage of seatbelts (Q196), [more] daily fruit intake (Q222), [less] diagnosed depression (Q107), and [better] sleep (Qs 22 and 24-26) with subjective feelings of [More] energy (Q58) and [intermediate to very good] self-assessment of health (Q54). In addition, healthy eating habits such as [higher] consumption of dairy products (Qs 226-228), [more] physical activity (Qs 12-17), and [more] time spent with friends and family (Qs 29, 30) are gathered into community 8. In contrast, unhealthy habits of eating [more] fast foods (Qs 198-204) are clustered alongside watching [more] television and movies (Qs 27, 28) in community 9. Furthermore, communities 12, 13, and 14 aggregate various non-communicable diseases with high burden of disease. Another intriguing finding is the division of questions related to symptoms generally associated with anxiety (community 5) and depressive (community 4) disorders into separate communities even though the questions were interspersed in the questionnaire.

**Figure 4.**
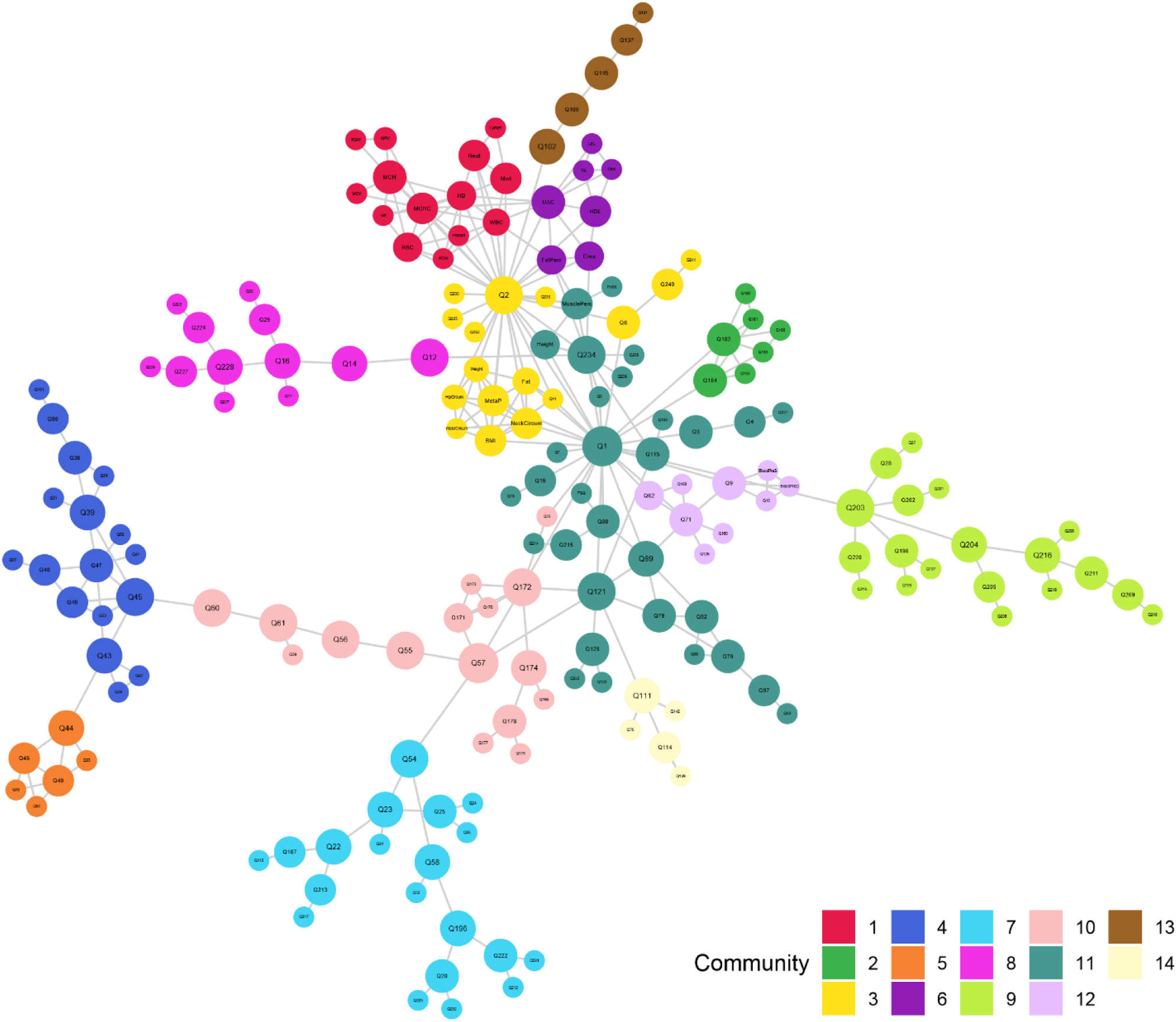
MF of the relationships between all variables in the dataset. Each node represents a variable and each edge indicates the presence of a relationship between a pair of variables based on mutual information. Nodes are colored based on their community (edge-dense clusters of interrelated variables) and their size indicates the logarithm of their betweenness centrality. A complete description of the variables and the communities to which they belong to is presented in Supplementary file 2.

It must be noted that the nature of the relationships which were indicated between brackets in the previous paragraph are not directly deducible from the minimal forest and were extracted upon further study of each connection (Figure S1). The complete description of detected communities and the variables clustered within them could be found in Supplementary file 2.

As was the case with the GGMs, individual connections in the minimal forest between pairs of variables may also hold valuable information. Even though many of the relationships identified here are well-known medical facts, some of the less studied associations are presented in Table 2.

**Table 2.**
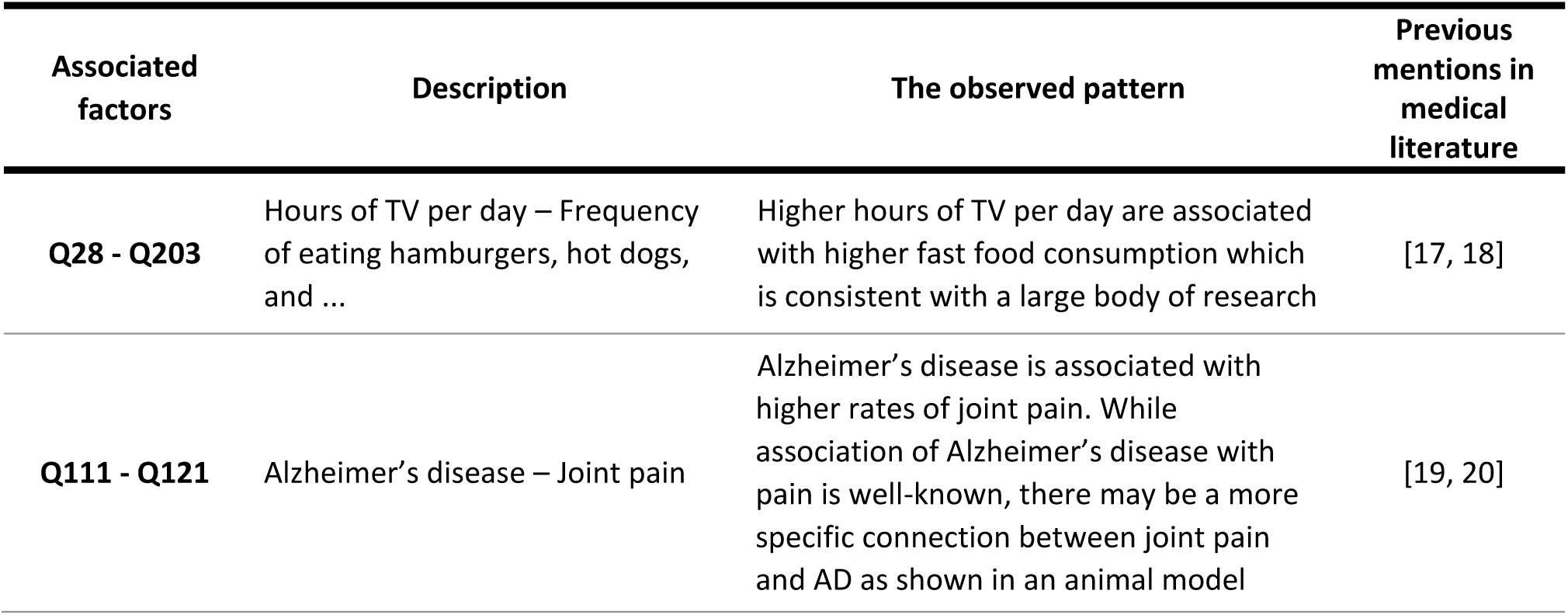
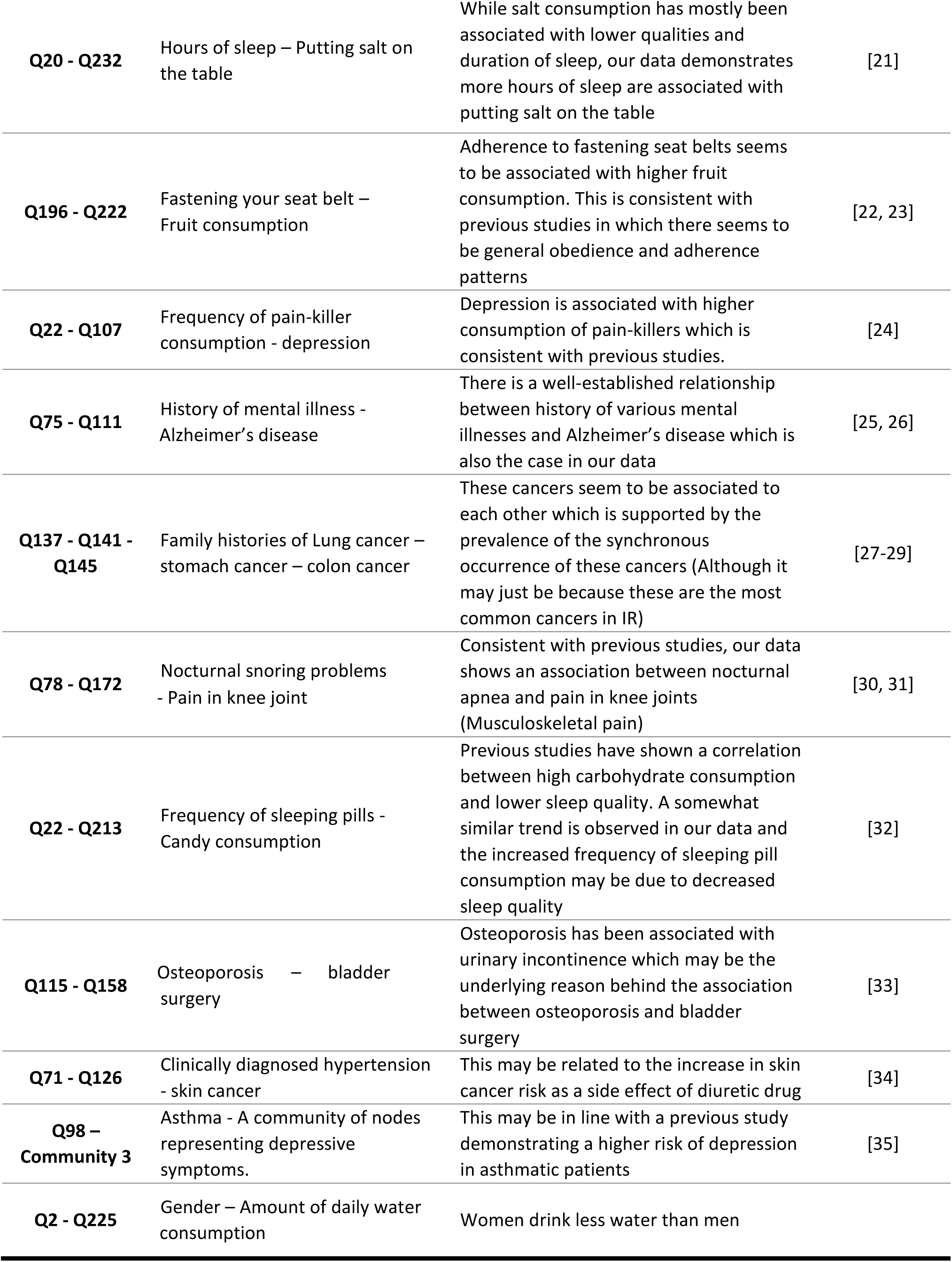
Interesting findings from minimal forest analysis of the YaHS dataset

As shown in Figure 4, there are three type of nodes within the graph with high values of betweenness centrality. These nodes are positioned in such a way that many of the shortest paths between variables (as explained in Methods) pass through them. The first group consists of nodes near the center of the graph and are, quite expectedly, age (Q1) and gender (Q2). The other group are nodes at the entry points of communities located towards the outer rim of the graph. Figuratively, these nodes act as gatekeepers of information (or influence) flow to-and-from their respective communities. They include: amount of physical pain experienced in a month (Q57) for community 10; subjective evaluation of health (Q54) for community 7; prevailing feeling of heart brokenness (Q45) for community 4; frequency of vigorous physical activity (Q12) for community 8; and frequency of eating hamburgers, pizzas, etc. (Q203) for community 9. Finally, the last group consists of nodes located between the central and outer nodes and, therefore, link them. This group consists of employment (Q234), joint pain (Q121), pain in knees (Q172), restriction in daily activities due to physical or mental problems (Q55, Q56), and amount of hurt felt due to physical and mental problems (Q60, Q61).

### 3.2 Deeper analysis of the identified relationships following general exploration

#### 3.2.1 VVKL analysis of blood cholesterol and HDL levels

It is obvious that an increase in blood cholesterol would be positively correlated to the increase in its HDL subfraction. However, what is more interesting is that some individuals have higher or lower HDL levels for a given blood cholesterol level. In other words, such individuals violate the expected linear trend between HDL and cholesterol. These variations have been attributed to both genetic [36] and environmental [37] factors.

Given the importance of cholesterol and its subfractions in human disease [38], identifying factors that may contribute to an increase in HDL is highly beneficial. These factors are presented in Figure 5. As was the case with GGM and MF, most of the findings by the VVKL method align with our current medical knowledge. Nevertheless, some interesting findings may also be observed.

**Figure 5.**
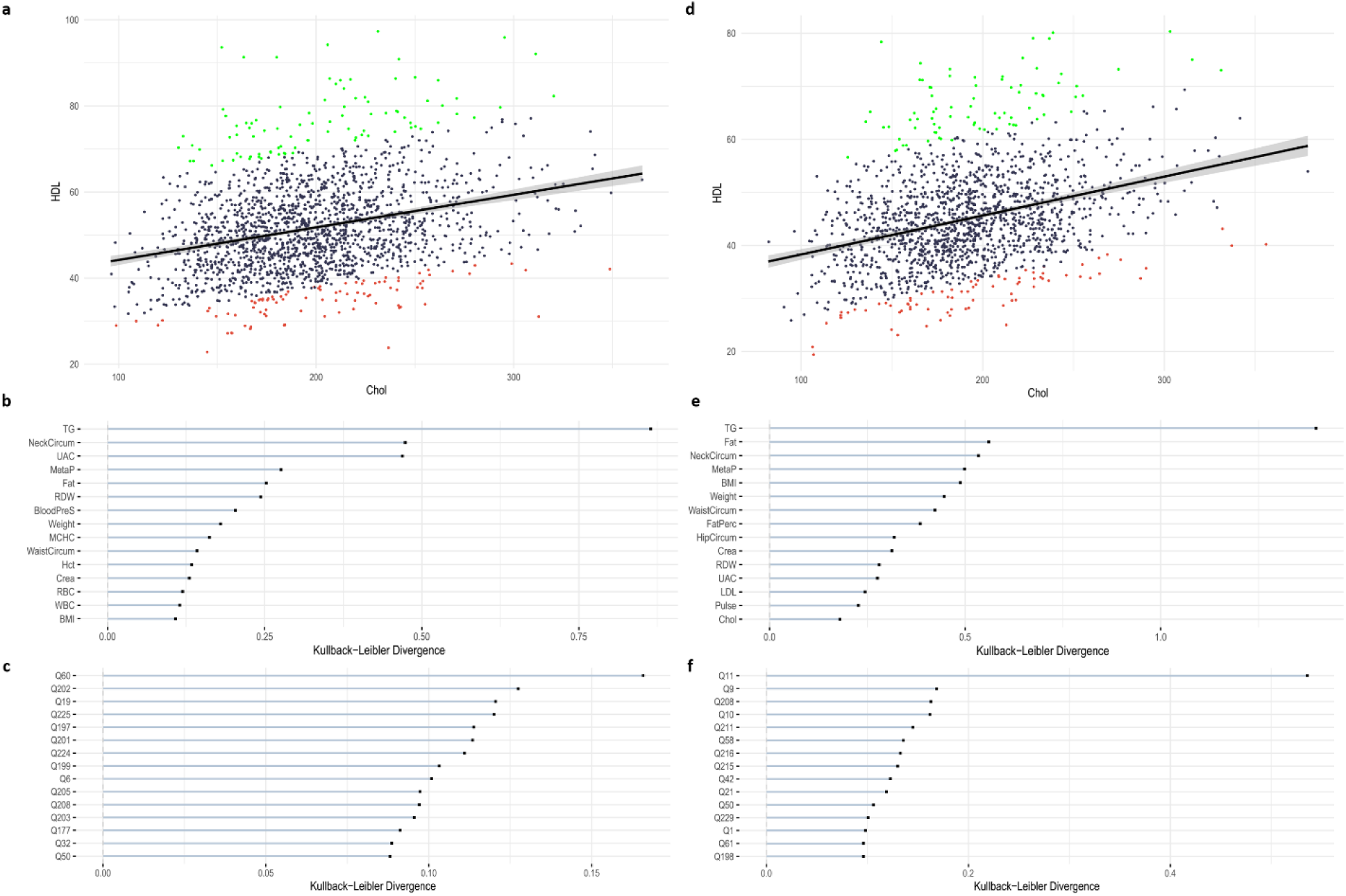
VVKL analysis on the association between total blood cholesterol (Chol) and blood high-density lipoprotein (HDL). **(a-c)** Plots showing the results of vvkl analysis in males. **(d-f)** Plots showing the results of vvkl analysis in females. **(a, d)** Scatter plot for Chol and HDL: Each blue point represents a sample. The data points violating the expected linear relationship between Chol and HDL are colored in green (up) and red (down). (**b, e)** KL divergences for continuous variables: The top 15 continuous variables with the highest KL values are shown in the bar plot for each gender. Each bar shows the scaled KL value. KL values are scaled such that each KL value is divided by the highest value among all KL values. **(c, f)** Similar to b and e but for categorical variables.

For example in continuous variables, Factors such as blood uric acid (UAC) [39], red blood cell distribution width (RDW) [40], body mass index (BMI) [41], hip circumference (hipCircum) [42, 43], and body fat percentage (FatPerc) [44] have well known associations with lipid profile. However, the findings from VVKL analysis suggest a difference in their relative importance between genders, which has also been previously mentioned for some of these variables [40, 42].

As for categorical variables, questions related to diet rank high in both genders. The results from VVKL analysis suggest that diet may have a stronger association than age and exercise with blood HDL levels. Furthermore, different aspects of diet have been emphasized in each gender. For example, egg consumption (Q208), bean consumption (Q211), type of oil used for cooking (Q216), and frequency of fried food consumption (Q215) are ranked high in males. In contrast, questions related the type of beverage drank with food (Q197), frequency of sweetened drink consumption (Q199), and removal of chicken skin (Q224) and meat fat (Q223) before cooking are have high KL divergence values in females. Furthermore, propensity to eat fast foods, as indicated by money spent on fast foods (Q202), reasons behind consuming fast foods (Q201), and frequency of eating hamburgers, pizzas, etc. (Q203), are generally more strongly associated with HDL levels in females. Importantly, all these factors have well known relationships with blood cholesterol and lipid profile [45-48] although gender differences within them may not be as clearly established.

Other significant differences between males and females include BMI as a categorical variable (Q11), which mirrors its continuous counterpart, and questions related to blood pressure (Q9 and Q10), which shiw a stronger association with blood HDL in men. Finally, mental conditions such as depression and anxiety (Q60) are very strongly related to HDL levels in females while lacking a significant association in males [49].

To further evaluate the performance of VKL, we also compared the two violating groups with Student’s t and χ^2^ tests. As shown in Table S3 and Table S4 (Supplementary file 2), the findings of vkl correspond nicely to that of the more traditional statistical tests.

### 2.4 Predictive multivariate elastic net models

We use the Elastic Net (EN) for predicting one categorical (osteoporosis) and one continuous (resting metabolic rate) variable based on all variables in the dataset. The factors selected by the EN as predictors are presented in Figure 6. Many of the variables selected by the EN as predictors have been shown to be related to the predicted condition as shown in Table S5and Table S6 (Supplementary file 2).

In Figure 6a, the predictions of the model built by the EN for osteoporosis are compared to results from question number 115 of the questionnaire. Positive responses to this question were only recorded if the condition was clinically diagnosed by dual X-ray absorptiometry (DEXA). The constructed model had an accuracy of 79 percent in predicting the condition. The receiver-operating curve for the model is presented in Figure S2 (Supplementary file 2). The model constructed by the EN had an area under the curve (AUC) of 87.4 percent and a sensitivity and specificity of 79 and 81 percent, respectively. Furthermore, the EN was used to construct a model for prediction of resting metabolic rate (MetaP in the dataset) compared to measurements by the Omron BF511 portable digital scale and body analyzer (Omron Inc. Nagoya, Japan) (Figure 6b). The predictions of the model had an R^2^ of 0.86 with the actual amounts for resting metabolic rate.

**Figure 6.**
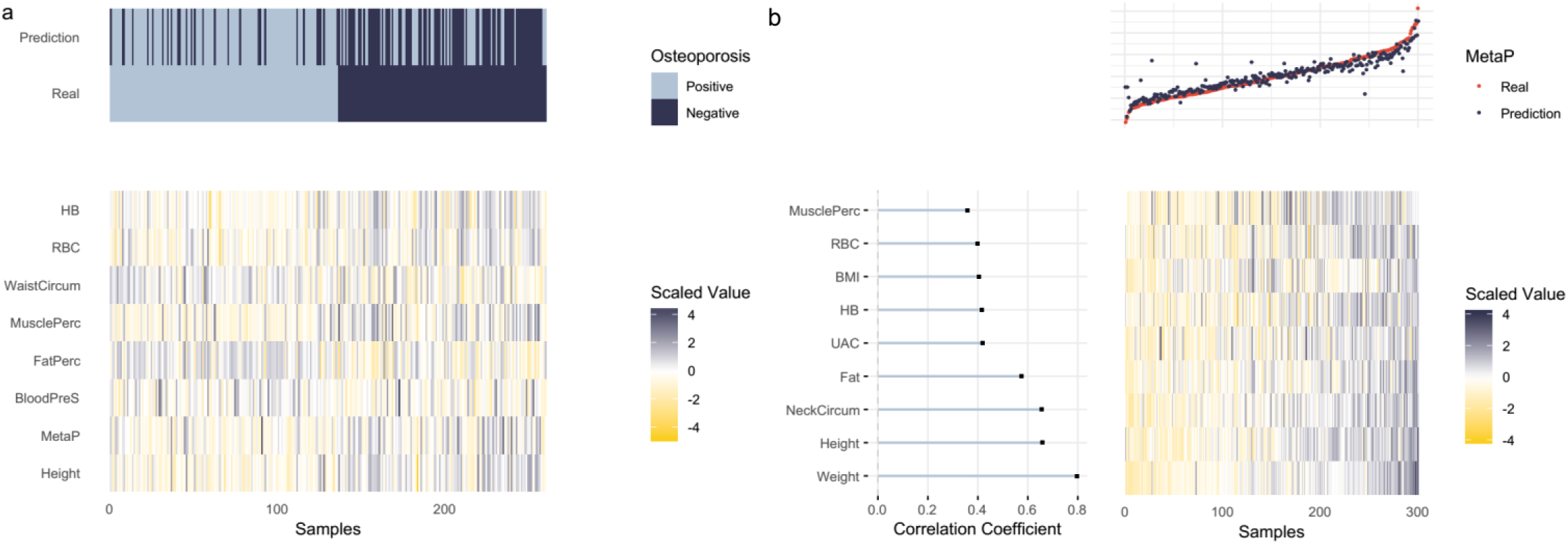
Multivariate elastic net model for categorical **(a)** and continuous **(b)** variables. The predictors with a non-zero coefficient in the model are shown. The scaled values of the predictors for a subsample of 100 participants are shown in the bottom heat map. **a)** Osteoporosis. The top heat map depicts the real values as well as the predicted values of the predicted variable, sorted for the value of the predicted variable. **b)** Resting metabolic rate. The correlation coefficients of the predictors and the predicted variable are shown as a horizontal bar plot on the left. A scatterplot showing the real values (red dots) and the predicted values (blue dots) of the predicted variable is illustrated at the top.

## 4 Discussion

In the MUVIS workflow, we propose general unbiased exploration as a means of determining all relevant relationships between all variables in the dataset. The large number of individuals and variables that are assessed in health surveys, along with the focus on studying a representative sample from the population, yield these datasets nicely to unbiased explorations. The issue with such exploration, however, is the sheer number of relationships that may be identified in large datasets, which would make interpretation almost impossible. The challenge, therefore, lies in extracting the most important and relevant associations, and then visualizing them in a practical way. In the following, we will discuss the roles of each method within the MUVIS pipeline and how to correctly interpret their results.

The first step in analyzing any dataset is examination of data quality. This quality control phase, which we have dubbed preprocessing in MUVIS, minimizes the unwanted confounding effects of factors related to data collection on the actual relationships between variables.

Following preprocessing, we use PGMs to estimate a sparse structure of connections among variables. The sparse nature of these graphical models means that they only show the statistically strongest, and therefore most relevant, relationships. As a result, one must bear in mind not all possible connections will be identified by these methods. PGMs have been used to this end in other areas of science as well, facilitating unbiased analysis in multivariate settings in bioinformatics [50, 51], studies of social networks [52, 53], and economics [54-56].

The fact that many of the relationships identified by the exploratory phases of the pipeline are in line with our current, comprehensively vetted, medical knowledge attest to the validity of these methods. This is evidently observed in the clustering of similar variables into communities with clear biomedical outlines in the GGM and MF methods. Furthermore, these methods are supported by the fact that many of the identified individual connections have previously been mentioned in the literature as demonstrated in Tables 1 and 2.

It must be noted that as in any other statistical method, any association identified by these algorithms requires further assessment and verification through checking the underlying data. For example, one caveat to some of the relationships identified for medically diagnosed diseases in our study is their low frequency in our population. This is especially true for cancers and mental conditions. As a case in point, it is difficult to comment on the robustness of the relationship between clinically diagnosed hypertension and skin cancer since our study population includes only 13 skin cancer patients, of which, 9 were hypertensive.

In addition to the identified relationships and communities, we propose a method for identifying keystone variables within the graphs using betweenness centrality (BC). This concept could have interesting implications. For example, if one intends to summarize a large survey into a smaller version while mostly preserving the amount of gathered information, the most information-rich variables may be good candidates for inclusion in the summarized version. As another example, one may consider a process of public health governance where it is necessary to achieve the highest efficacy despite the lowest expenditure of resources. Therefore, policy makers may use this method to identify the variables which could have the largest effects if manipulated. In addition, since all variables within a graph are ranked based on BC, the policy makers would have flexibility in striking a balance between choosing the most influential variables and the limitations they may face, most notably, cost and feasibility.

Following the identification and validation of relationships, the VVKL method enables the researcher to delve deeper into the connections and how they are influenced by other variables in the dataset. VVKL identifies the data points that violate the expected linear relationship between two variables of interest and divides them into two groups, upward and downward violators. Then, it explores the rest of the dataset to find variables that are differently distributed between the two groups. Not only does this method help in gaining a better understanding of the association between two variables of interest, it is also able to find new connections between the variables of interest and the variables differentially distributed between the violator groups. Therefore, VVKL is another unbiased exploratory step in the MUVIS workflow.

Indeed, as shown in Tables S3 and S4 (Supplementary file 2), it is also possible to compare the two violator groups using the traditional statistical methods of Student’s t and χ^2^ tests. However, unlike these methods, VKL analysis is applicable on both categorical and continuous variables and readily provides an effect size (KL divergence) to gauge the significance of the associations it finds. Given the general consistency of VKL’s results with that of Student’s t and χ^2^ tests, we propose VVKL as a powerful and robust alternative which is able to simultaneously analyze and rank the influence of all variables in a dataset on a desired linear relationship.

Finally, the Elastic Net (EN) algorithm provides an effective and simple to use method for constructing predictive models in medicine. In an era with ballooning medical costs and increasing healthcare discrepancies, construction of models using relatively accessible medical features to predict more complex and more expensively diagnosed medical conditions is necessary. As was the case with BC values in the PGMs, the EN provides a flexible framework for model construction as it ranks all variables in the dataset in terms of their predictive value for our variable of interest. As a result, it becomes possible to find a balance between model accuracy and any factor, such as cost, that may limit the number and nature of predictors. Importantly, since the elastic net model surveys all variables in the database, it may select ones that may not have been readily hypothesized to be useful predictors, corresponding to the general unbiased exploratory approach of MUVIS.

In conclusion, the MUVIS workflow is an easy to use and effective tool for extracting and visualizing meaningful information from large datasets and is able to do so by offering the following solutions to the challenges presented in the Introduction section:

1. The methods proposed in this pipeline (MF, VVKL) are applicable simultaneously on continuous and categorical variables. Therefore, it becomes possible to directly compare these different types of variables within a single statistical framework.
2. Powerful quality control methods in the preprocessing phase ensure data quality and lack of confounding effects by elements not related to the variables themselves.
3. The unbiased nature of all methods in MUVIS minimize the effects of biases in study design and data analysis.
4. As a solution to the problem with “hyper-significant” p-values, we propose using partial correlation, mutual information, and Kullback-Leibler (KL) divergence between distributions of variables as indicators of the strengths of relationships between variables.
5. For construction of predictive models, we propose using the EN method since it is both unbiased and well suited for handling large datasets.

In this study, we intentionally used data from a general health survey which contained all the most common and extensively studied health indices. This enabled us to validate the relationships identified within our study with an extensive body of scientific literature. Another such analysis in Supplementary file 1 presents findings from the application of MUVIS on the National Health and Nutrition Examination Survey (NHANES) dataset. Another reason behind choosing the Yazd Health Survey as our data source was its repetition in 5-year intervals. Therefore, the associations and predictions proposed by MUVIS could be further validated through time.

A consequence of this approach was that most relationships identified in this study were not novel and had been previously mentioned. Nevertheless, as a future step, application of MUVIS on general health surveys in other populations may find interesting cross-population differences. Furthermore, we believe there is clear potential for finding more novel and interesting associations if MUVIS is applied on more condition-oriented datasets. Therefore, another step is to apply this workflow on datasets with higher prevalence of various medical conditions. Finally, there is great room for improvement for the MUVIS package including, but not limited to, devising novel methods for identification of information-rich variables, addition of other high-accuracy predictive models, increase of computational efficiency and speed of its methods, and reduction of it dependency on other R packages to improve its ease of use.

## Data Availability

- The source code for the MUVIS package in R:
https://github.com/bAIo-lab/muvis
- The source code for the application of MUVIS on the YaHS dataset: https://github.com/vdblm/YaHS
- The YaHS questionnaire:
http://www.yahs-ziba.com/index.php/colaboration/study-catalogue

## Acknowledgements

We gratefully acknowledge the creative comments of Alireza Modir-Shanechi, Shahryar Javidi, and Amirreza Asadzadeh on information-theoretic interpretation of representative selection (‘information rich’ nodes) from graph communities. Authors would also like to thank the Yazd people for participation in the study, graduate students for participation in study design, and the workers and managers of Yazd’s health centers for acquiring the data.

## Author contributions statement

E.H. proposed the workflow, and implemented it in R language. E.H. and V.B. developed the MUVIS package, performed the statistical analyses, and visualized the results. M.A.S. interpreted the results, suggested modifications to the methods in MUVIS to improve their clinical relevance, proposed the utility of identifying information-rich variables, and wrote the manuscript. A.S reviewed the results and suggested modifications to the analysis workflow. N.A. coordinated the study and reviewed the results. M.M. and M.S. provided the data, reviewed the results, and proposed the question which led to developing VVKL. E.H., M.M., and A.S. revised the manuscript. A.S. supervised the project.

## Code and Data Availability

The source code for the MUVIS package in R: https://github.com/bAIo-lab/muvis

The source code for the application of MUVIS on the YaHS dataset: https://github.com/vdblm/YaHS

The YaHS questionnaire: http://www.yahs-ziba.com/index.php/colaboration/study-catalogue

Data from subjects 1 - 100 is presented as a sample from the YaHS dataset in Supplementary file 3.

